# Old tools, new applications: use of environmental bacteriophages for typhoid surveillance and evaluating vaccine impact

**DOI:** 10.1101/2023.02.14.23285884

**Authors:** Yogesh Hooda, Shuborno Islam, Rathin Kabiraj, Hafizur Rahman, Kesia E. da Silva, Rajan Saha Raju, Stephen P Luby, Jason R Andrews, Samir K Saha, Senjuti Saha

**Author notes:** Correspondence to Senjuti Saha, Director & Scientist, Child Health Research Foundation, Dhaka, Bangladesh.

## Abstract

Typhoid-conjugate vaccines (TCVs) provide an opportunity to reduce the burden of typhoid fever, caused by *Salmonella* Typhi, in endemic areas. As policymakers design vaccination strategies, accurate and high-resolution data on disease burden is crucial. However, traditional blood culture-based surveillance is resource-extensive, prohibiting its large-scale and sustainable implementation. *Salmonella* Typhi is a water-borne pathogen, and here, we tested the potential of Typhi-specific bacteriophage surveillance in surface water bodies as a low-cost tool to identify where *Salmonella* Typhi circulates in the environment. In 2021, water samples were collected and tested for the presence of *Salmonella* Typhi bacteriophages at two sites in Bangladesh: urban capital city, Dhaka, and a rural district, Mirzapur. *Salmonella* Typhi-specific bacteriophages were detected in 66 of 211 (31%) environmental samples in Dhaka, in comparison to 3 of 92 (3%) environmental samples from Mirzapur. In the same year, 4,620 blood cultures at the two largest pediatric hospitals of Dhaka yielded 215 (5%) culture-confirmed typhoid cases, and 3,788 blood cultures in the largest hospital of Mirzapur yielded 2 (0.05%) cases. 75% (52/69) of positive phage samples were collected from sewage. All isolated phages were tested against a panel of isolates from different *Salmonella* Typhi genotypes circulating in Bangladesh and were found to exhibit a diverse killing spectrum, indicating diverse bacteriophages were isolated. These results suggest an association between the presence of Typhi-specific phages in the environment and the burden of typhoid fever, and the potential of utilizing environmental phage surveillance as a low-cost tool to assist policy decisions on typhoid control.

**Highlights:**

- Typhi-specific bacteriophages can be isolated from surface waters in endemic countries using low-cost methods
- More Typhi-specific bacteriophages are obtained in areas with higher typhoid cases
- Typhi-specific bacteriophages exhibit diverse activity spectrum against a panel of *Salmonella* Typhi isolates circulating in Bangladesh
- Environmental surveillance can be used as a tool to predict typhoid burden

**Graphical abstract:** 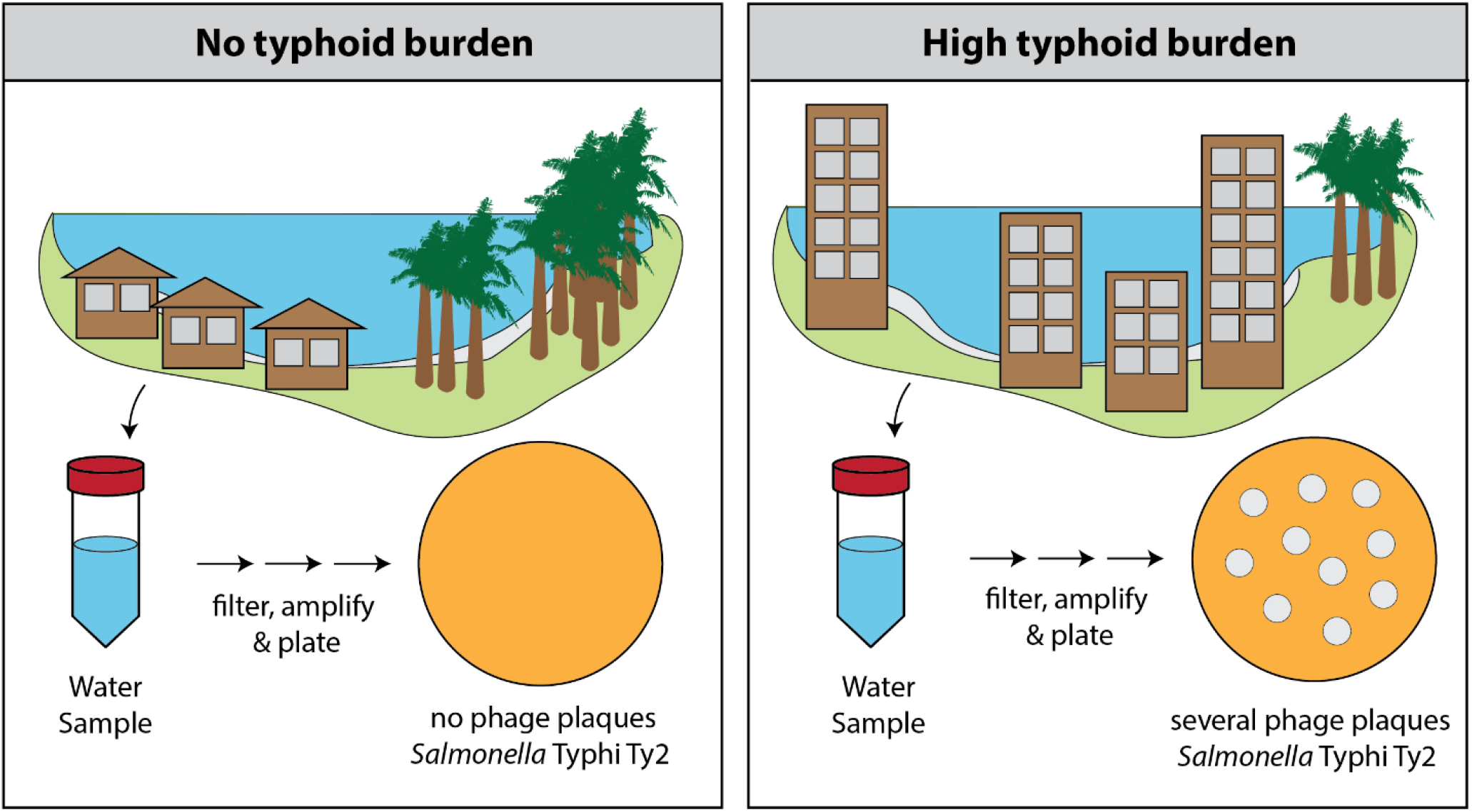

## INTRODUCTION

Typhoid fever is a systemic infection caused by the water-borne pathogen *Salmonella* enterica serovar Typhi. This pathogen is common in many low- and middle-income countries (LMICs) causing an estimated 135 000 deaths and 14 million infections globally (Stanaway et al., 2019). The World Health Organization (WHO) recommended the use of typhoid conjugate vaccines in settings with high burden of typhoid fever (Andrews et al., 2018; Saha et al., 2021). Countries have begun implementing these suggestions, and several countries have performed large-scale clinical trials to demonstrate that these vaccines exhibit 80-85% efficacy in preventing infections (Patel et al., 2021; Qadri et al., 2021; Shakya et al., 2019). Typhoid-conjugate vaccines are currently being utilize to tackle an extensively-drug resistant (XDR) *Salmonella* Typhi outbreak in Hyderabad, Pakistan, where vaccine effectiveness has been demonstrated to be high (Batool et al., 2021).

Decisions are currently being made to roll out typhoid-conjugate vaccines in other endemic countries, however, accurate and high-resolution spatial data regarding the burden of typhoid is required for optimal use of the available vaccines. The current estimates of burden of typhoid fever in countries have primarily come from modelling studies based on limited surveillance data and do not provide the geographical and temporal resolution within local communities and countries to design effective preventive and treatment measures. The paucity of the data is in large part because traditional blood culture surveillance is resource extensive, requires clinical laboratory infrastructure and trained health and research professionals. Consequently, very few LMICs routinely conduct these studies and a few that do, tend to focus on high-risk urban settings, and are not able to sustain these studies as a regular part of health infrastructure. This has led to search for low-cost and sustainable methods to supplement traditional clinical surveillance systems (Andrews et al., 2020; Saha et al., 2019). To this end, environmental surveillance strategies that can identify *Salmonella* Typhi in different water supply have been proposed. In recent years, environmental wastewater surveillance has been used for early detection and monitoring the spread of SARS-CoV-2 (Karthikeyan et al., 2022, 2021) and poliovirus (Asghar et al., 2014) among others.

Previous studies have shown that high detectable levels of *Salmonella* Typhi in the water supply overlaps with areas of disease burden, suggesting sampling water could be utilized as a preliminary surveillance proxy (Andrews et al., 2020; Sikorski and Levine, 2020). While it has not been possible to reproducibly culture *Salmonella* Typhi directly from environmental water, quantitative polymerase chain reaction (qPCR) has been proposed to detect *Salmonella* Typhi DNA (Liu et al., 2021; Saha et al., 2019). However, qPCR-based methods cannot be replicated in most settings with typhoid burden, due to the lack of infrastructure (machines, molecular techniques etc.) and the high associated cost. To address this gap, we investigated if the presence of bacteriophages in the environmental water samples could be used as a proxy to estimate the prevalence of *Salmonella* Typhi in water bodies.

Bacteriophages (or phages) are viruses that infect bacteria and bacterial abundance, phenotypic characteristics, and long-term evolutionary trajectory. Bacteriophages are very specific to their host bacterial species and often able to discriminate between different sub-populations based on minor genetic differences in host receptor and surface epitopes (De Smet et al., 2017). Bacteriophages against *Salmonella* Typhi (Typhi phages) were first reported in the 1940’s (Felix, 1955). Typhi phages were extensively used for bacterial typing in the 1940s-80s before the advent of molecular diagnostic methods such as PCR and genomic sequencing (Farmer et al., 1975). However, little has been in done in the last 50 years and there is a lack of contemporary literature regarding Typhi phages from typhoid-endemic countries. This motivated us to initiate a pilot study to determine the feasibility of detecting *Salmonella* Typhi-specific phages in water bodies and typhoid burden at two geographic regions: Dhaka, a city of 9 million people with a high typhoid burden and Mirzapur, a rural district with 340,000 people that has low typhoid burden. Overall, our work provides a pilot study for testing if the prevalence of phages correlates with local typhoid burden.

## METHODS

### Data from clinical surveillance and ethical considerations

The clinical laboratories of Bangladesh Shishu Hospital and Institute [BSHI], Shishu Shasthya Foundation Hospital, [SSFH]) and Kumudini Women Medical College and Hospital [KMWCH] are part of the laboratory network of Child Health Research Foundation CHRF, where all data are stored electronically. These laboratories are part of the WHO-supported Invasive Bacterial Vaccine Preventable Surveillance conducted at the CHRF and has been described earlier (Saha et al., 2017). Data on blood culture surveillance was obtained from these electronic records. The protocols for data use were approved by the ethics review committees of the Bangladesh Institute of Child Health, BSHI. Blood samples were collected and received at the laboratories as part of routine clinical care.

### Environmental sample collection and bacteriophage isolation

Water samples were collected from sewage drains, rivers, ponds, lakes, and stagnant water bodies selected based on accessibility. Stagnant waters are defined as temporary water bodies that form after flooding or rainfall that cannot be used for livestock or human use, often due to poor water quality. A sterile cup, attached to a rope, was used to collect >10 ml of water sample from each source. Maintaining sterile techniques, the sample was transferred into a sterile bottle for transportation. Ten ml of the sample was centrifuged at 10,000 rpm for 5 minutes in a 15 mL conical tube to pellet debris and bacteria or larger microbes. The supernatant was passed through a 0.22 µm PES syringe filter into a new tube. The filtered sample was stored at 4&C up to 72 hours before use for further experiments.

A total of 500 µL of the filtered water sample was mixed with 450 µl of LB broth and 50 µl of overnight liquid Ty2 host culture in a 2 mL microcentrifuge tube. This mixture was incubated at 37&C for 2 hours followed by the addition of 2-3 drops of chloroform to lyse and kill all bacteria in the mixture without affecting bacteriophages. The mixture was then centrifuged at 10,000 rpm for 10 minutes and 750 µL of the supernatant potentially containing enriched *Salmonella* Typhi bacteriophages was transferred to a new tube.

### Bacteriophage detection & propagation

For bacteriophage detection, the enriched sample was tested using the double-layer agar method described earlier (Kropinski et al., 2009). In brief, 100 µl of the sample was incubated with 200 µl of overnight culture host strain Ty2 for 20 minutes. The entire 300 µl and was mixed with 3 mL of molten soft agar (0.7% agar) and poured over solid hard agar plates (1.5% agar). These plates were incubated at 37&C for 14-16 hours and the appearance of plaques the next day indicated the presence of bacteriophages in the sample. For each plaque morphology, a plaque was picked using a 100 µl pipette tip and resuspended in 100 µl of tryptic soy broth. Two drops of chloroform were added to the resuspension followed by 10 minutes of incubation, and a final centrifugation at 10,000 rpm for 10 minutes. 70 µl of the supernatant was transferred to a new tube which contained a clone of phages of a single morphology.

### Activity spectra of Typhi phages

To confirm that these phages are Typhi specific, 3 µl dilutions of isolated phages were spotted on strains of *Salmonella* enteric serovars Paratyphi A (E321: 1100582310), Paratyphi B (366817) and Typhimurium LT2, and Gram-negative bacteria *Escherichia coli* (ATCC: 25922), *Klebsiella pneumoniae* (ATCC: 700603 & ATCC: 9349) and Pseudomonas aeruginosa (ATCC: 27853) using the double layer methodology.

To test diversity of the phages, they were also spotted on 14 *Salmonella* Typhi isolates collected from blood culture belonging to different genotypes previously described to be present in Bangladesh (Tanmoy et al., 2018). The plates were incubated overnight at 37&C for 14-16 hours and the plates visualized for zones of clearing. The killing activities against the different genotypes were recorded and hierarchical clustering and heat map generation was made using the R package *heatmaply*.

## Results

### Clinical typhoid surveillance

Since 2012, we have been conducting surveillance to monitor enteric fever, pneumonia, meningitis, and sepsis (as part of the Invasive Bacterial Vaccine Preventable Disease Surveillance of the WHO) in two hospitals in urban Dhaka (BSHI and SSFH), and in one hospital in Mirzapur (KMWCH), a rural district approximately 60 km north of Dhaka (Saha et al., 2018, 2017). In 2021, we performed 4,620 blood cultures in BSHI & SSFH, of which 215 (4.7%) were culture-confirmed for *Salmonella* Typhi. In contrast, at KMWCH, during the same period 3,788 blood cultures were performed and only 2 (0.05%) were culture-confirmed for *Salmonella* Typhi (Table 1). The burden of culture-confirmed cases of typhoid fever in Mirzapur in 100-fold lower.

**Table 1.** Blood culture positivity of *Salmonella* Typhi in clinical surveillance, and bacteriophage positivity in environmental water samples in Dhaka and Mirzapur in Bangladesh.

| Study site in Bangladesh | Type of surveillance | Type of sample | Total no. of samples tested | No. of *Salmonella* Typhi/ phage positive sample | % Positive samples |
| --- | --- | --- | --- | --- | --- |
| Dhaka | Clinical surveillance | Blood | 4620 | 215 | 5% |
|  | Environmental surveillance | Sewage | 168 | 51 | 30% |
|  |  | Lake | 18 | 10 | 56% |
|  |  | Pond | 15 | 2 | 13% |
|  |  | River | 5 | 2 | 40% |
|  |  | Stagnant water | 5 | 1 | 20% |
|  |  | Total | 211 | 66 | 31% |
| Mirzapur | Clinical surveillance | Blood | 3788 | 2 | 0.05% |
|  | Environmental surveillance | Sewage | 3 | 1 | 33% |
|  |  | Pond | 48 | 0 | 0% |
|  |  | River | 4 | 0 | 0% |
|  |  | Stagnant water | 37 | 2 | 5% |
|  |  | Total | 92 | 3 | 3% |

### Environmental phage surveillance

Between August 2021 and December 2021, we collected 211 environmental water samples in the Dhaka region. These samples constituted of sewage water (n = 168), lake (n = 18), pond (n = 15), river (n = 5) and stagnant water (n = 5) from different sites across Dhaka (Figure 1A). Sixty-six of the 211 samples (31%) exhibited plaque formation, and in all cases, the plaques could be propagated confirming the presence of active bacteriophages. We observed at least two morphologies in 16 samples suggesting different phages in the same sample. Phages of different morphologies could be further purified during propagation bringing the total number isolated lytic phages to 82 from 211 samples in Dhaka. The distribution of positive and negative samples showed that Typhi phages were present in all types of water bodies tested (Table 1).

**Figure 1:**
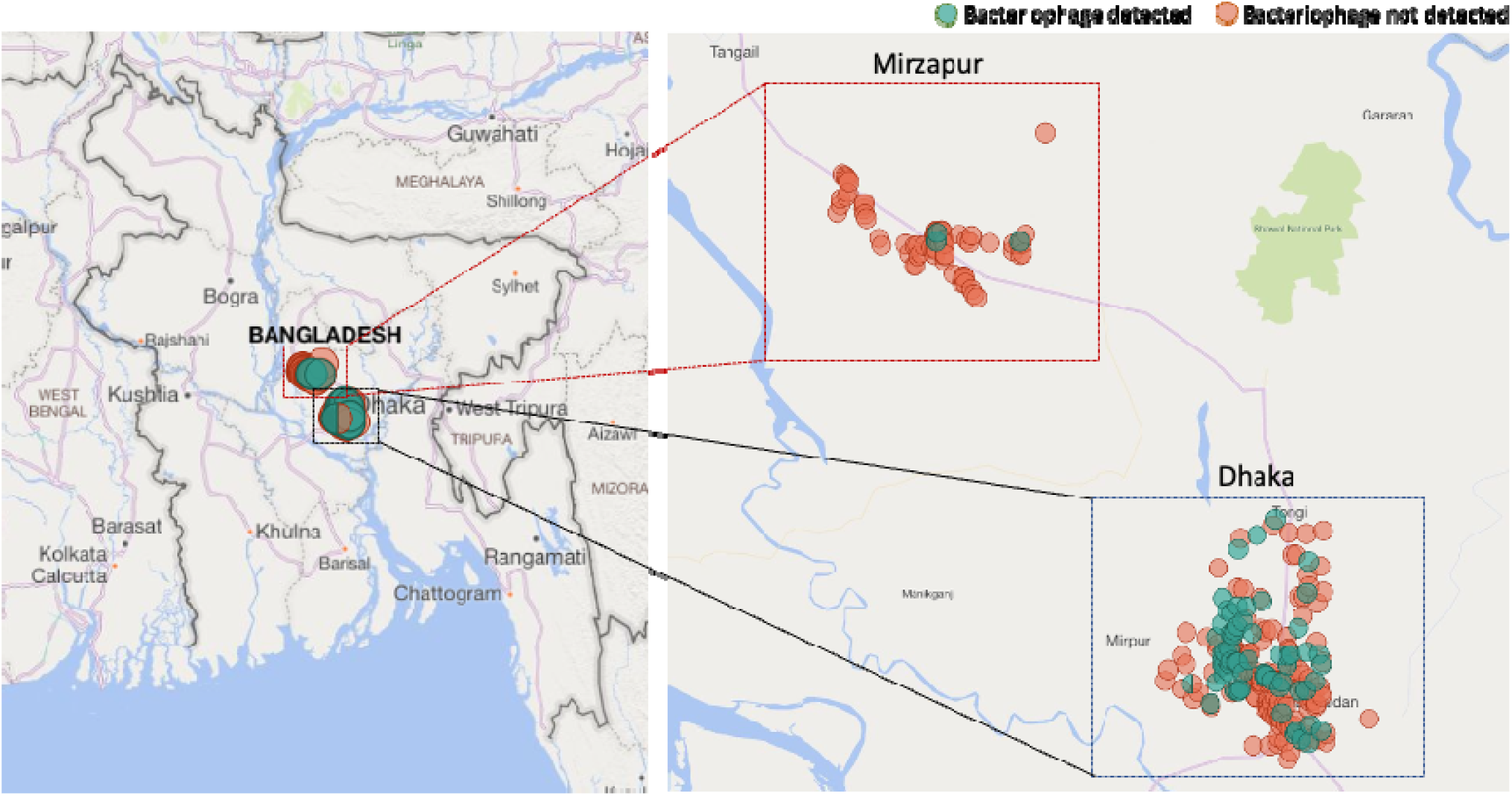
Location of water sample collection and detection of *Salmonella* Typhi phages in water samples of urban Dhaka and rural Mirzapur, Bangladesh.

In contrast, a total of 92 environmental samples were collected during the same time from the Mirzapur region (Figure 1B). These samples constituted of sewage water (n = 3), pond (n = 48), river (n = 4), and stagnant water (n = 37). A total of 3 samples showed positive phage lytic activity in the 92 samples tested (3%), the details of which are provided in Table 1. Two different plaque morphologies were noted in one sample, bringing a total of 4 isolated phages from these 92 samples (Table S1). No phages were detected in ponds or river; one positive sample was from sewage water (n=1, 33%) and two from stagnant water. Overall, phage prevalence in water bodies in Dhaka (31%) was 10-fold higher than water bodies in Mirzapur (3%), correlating with the culture-confirmed typhoid cases in the largest hospitals of the regions.

genotypes (X-axis) circulating in Bangladesh. Blue represents activity/plaque formation, white represents no activity/no plaque formation. The phages on the Y-axis are labelled based on the site of isolation. Phages from Mirzapur are labelled as MZ, and From Dhaka are labelled as DK. Multiple phages isolated from the sample are indicated with .1 and .2 at the end of the label. The heat map was made using the R package *heatmaply*.

### Host range and diversity of Typhi phages

To test the specificity of phages isolated, we tested all 86 isolated phages against closely related *Salmonella* enteric serovars Typhimurium, Paratyphi A and Paratyphi B, and closely related *Enterobacteriaceae* species Escherichia coli and *Klebsiella pneumoniae*. Another gamma-proteobacterium *Pseudomonas aeruginosa* was also included. No cross activity was observed, depicting that these phages are highly specific to *Salmonella* Typhi.

Furthermore, to understand the diversity of the phages collected, we tested the killing spectrum of the 86 phages against a panel of 17 *Salmonella* Typhi strains each representing a different genotype circulating in Bangladesh (Tanmoy et al., 2018) (Figure 2). The phages showed a diverse killing spectrum and could be grouped into 48 clusters based on their killing activity. This demonstrated that the circulating *Salmonella* Typhi strains in Bangladesh are not equally susceptible to all environmental phages; some are more susceptible than others. In addition, phages with different plaque morphologies from the same samples often displayed (for example DK_23.1 and DK_23.2) different spectra, indicating that multiple phages can circulate in the same water body at the same time.

**Figure 2:**
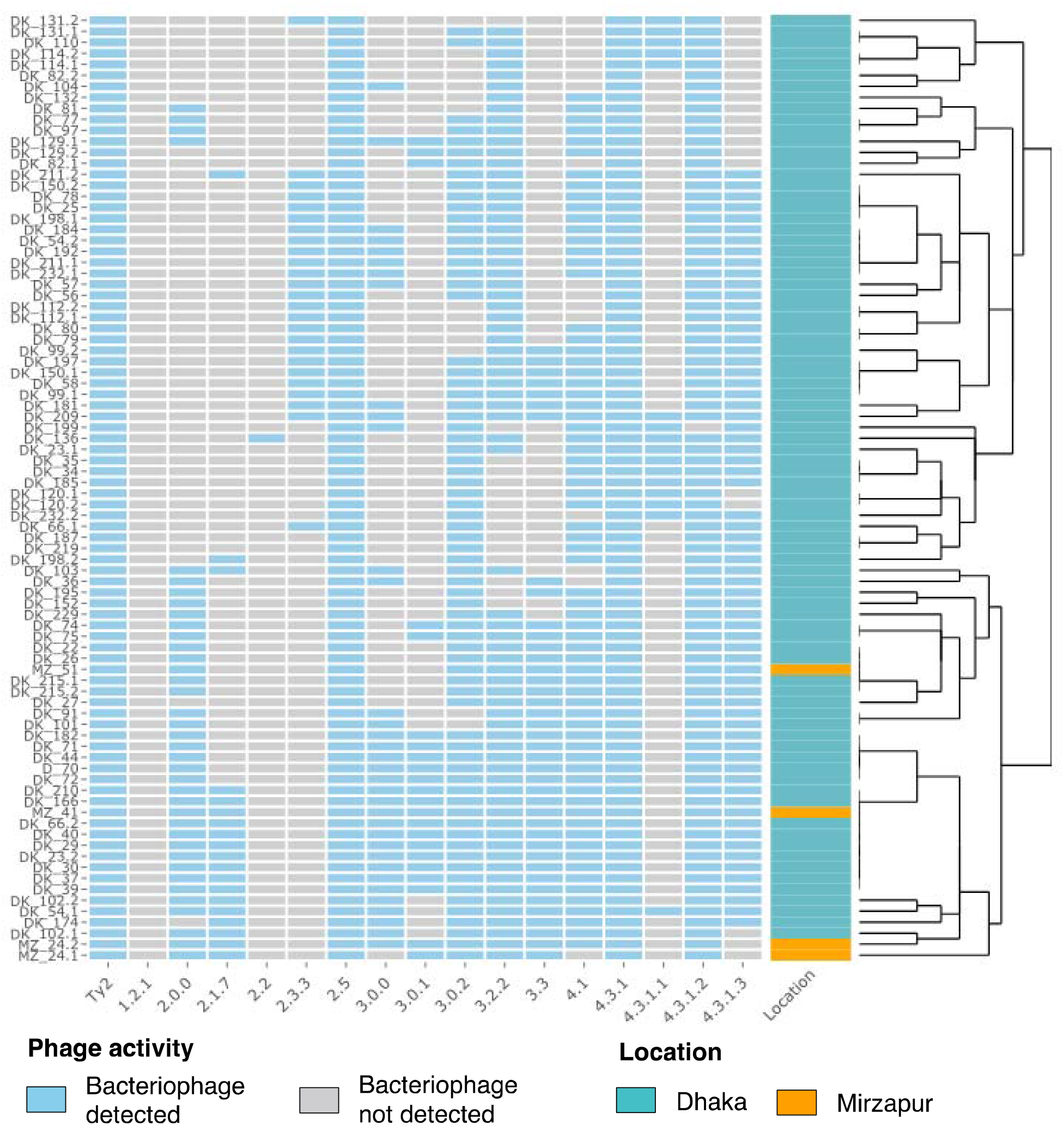
Hierarchical clustering of the activity spectra of the 86 bacteriophages isolated in the study. All isolated phages were spotted on Salmonella Typhi isolates belonging to different.

## Discussion

Our pilot study shows that detection of bacteriophages specific to *Salmonella* Typhi may be a rapid environmental surveillance method to understand the presence of typhoid fever in the community. 33% of environmental samples collected in Dhaka contained phages, where blood culture positivity was 5%; by comparison, 3% of environmental samples collected in Mirzapur contained phages, where blood culture positivity was 0.05%. Of all the sources of water collected, 52/69 (75%) of samples were collected from sewage indicating that wastewater surveillance is well-suited for monitoring typhoid fever. In Dhaka, a city with high burden of typhoid fever, high phage positivity was also seen in water samples collected from lakes (56%) and rivers (40%). Similar results were obtained from the related study from Nepal, another typhoid endemic country in South Asia.

Given the minimal resources required for undertaking environmental phage surveillance, it can be readily rolled out in resource-constrained settings and can complement existing surveillance strategies. Sample processing required minimal resources, which primarily include collection bottles and/or tubes, syringes with 0.22 syringe filters, petri dishes, media for bacterial growth, an incubator, a centrifuge, pipettes, and the laboratory strain Ty2 of *Salmonella* Typhi (See Methods). The cost for consumables for each sample less than USD 10 in Bangladesh, this might vary slightly based on location. Such low costs of sample collection and easy processing means that phage surveillance is more scalable than PCR-based molecular methods, which are both resource and expertise intensive. Furthermore, the stability of phages (Jończyk et al., 2011) in water means that samples can be collected from all over the country and tested at any location with minimal fears of loss of quality of the results obtained. In contrast, environmental DNA degrades fast, and thus PCR based surveillance typically requires transport of refrigerated or frozen samples.

Limited work has been done in investigating the role that phages play in the ecology of *Salmonella* Typhi. The abundance and diversity of Typhi phages in terms of the killing spectrum (seen in our study) and the genetic sequences (seen in related study from Nepal) highlight that Typhi bacteriophages are likely to play an important role in determining the spread and seasonality of *Salmonella* Typhi. Additionally, certain genotypes of *Salmonella* Typhi (such as 1.2.1, 2.2 and 4.3.1.1) are more resistant to phages vs others (such as 4.3.1.2, 4.3.1 and 2.5). The molecular basis of the observed differences in phage resistance amongst different genotypes may be due to differences in receptor sequences and/or modifications, or presence of phage defense systems (such as CRISPR-Cas). Future studies addressing these questions may be helpful in determining the impact phages have on circulating *Salmonella* Typhi population (Faruque et al., 2005; LeGault et al., 2021). Additionally, with rising antimicrobial drug resistance, renewed research on Typhi phages might provide alternate weapons for clinical development.

The results in this study should be interpreted within the context of the following limitations. First, the number of samples obtained does not fully represent the number of water bodies present in Dhaka or Mirzapur. Second, no water sampling could be conducted from July-August during the study period due to monsoon-associated floods. Third, all phage amplification steps were done in *Salmonella* Typhi strain Ty2, and hence phages that do not infect this strain were missed. Fourth, sewage samples were underrepresented in Mirzapur due to lack of a sewage system in many parts of rural areas. Finally, comparison with the PCR-based assays will be required to identify the specificity and sensitivity of phage detection. Expansion of this study to other regions of Bangladesh where clinical data is available will help in resolving some of these limitations. It would also be helpful to examine temporal trends in phage positivity and how these trends correlate with the seasonality of typhoid fever.

In summary, in this study, we propose a simple, cost-effective, and scalable method for conducting environmental surveillance for typhoid fever. The method uses standard microbiological laboratory infrastructure and techniques to detect Typhi-specific bacteriophages. Environmental phage surveillance can be used to estimate typhoid in other countries, including in Sub-Saharan Africa where limited epidemiological data on typhoid fever is available (Carey and Steele, 2019; Kim et al., 2022). Environmental phage surveillance may be also applied to map routes of disease transmission by focusing on wastewater/sewage facilities in the region. This tool, combined with traditional blood culture surveillance (Wang et al., 2020), can generate community-level data to evaluate the impact of interventions including the introduction of TCV, water improvement projects, and sanitation and hygiene systems.

## Data Availability

All data produced in the present study are available upon request to the corresponding author: Dr. Senjuti Saha

